# Evaluating the real-world performance of plasma pTau217 and pTau181 in a Southeast Asian tertiary memory clinic

**DOI:** 10.1101/2025.05.22.25328192

**Authors:** Yi Jayne Tan, Raziyeh Mohammadi, Seyed Ehsan Saffari, Nicole Isabella Tan, Louis SS Lim, Shermine HL Low, Louis CS Tan, Hui Jin Chiew, Kok Pin Ng, Shahul Hameed, Simon KS Ting, Adeline SL Ng

**Author notes:** **Corresponding author:** Assoc Prof Adeline SL Ng, Department of Neurology, National Neuroscience Institute, 11 Jalan Tan Tock Seng, Singapore 308433.

## Abstract

**Background:** Recent advancements in plasma pTau217 have shown its potential as a reliable biomarker for detecting brain amyloid pathology (Aβ) in Alzheimer’s disease (AD). However, its application in real-world settings remains limited, particularly in diverse populations such as those in Asia. While plasma pTau217 has demonstrated high accuracy in research cohorts, most studies have been conducted in Western populations. We evaluated the performance of plasma pTau217 and pTau181 with longitudinal cognition in a real-world tertiary memory clinic setting.

**Methods:** Plasma ptau217 was measured using ALZpath SIMOA assay in 291 participants [68 healthy controls (HC), 100 mild cognitive impairment (MCI), 86 AD, 37 non-AD dementia] from National Neuroscience Institute, Singapore. Performances of pTau217, derived reference ranges (low-, intermediate-, high-) for amyloid positivity and associations with cognitive decline over 4 years was evaluated.

**Results:** Plasma pTau217 [(area under the curve (AUC) = 0.942)] outperformed pTau181 (AUC= 0.909) in differentiating AD from other disease groups. Amongst patients with known amyloid status (n=40), pTau217 identified Aβ+ with AUC= 0.961 vs pTau181 with AUC= 0.917. Using a two cut-point approach, a 95% sensitivity and specificity for Aβ+ corresponded to pTau217 >0.66 pg/ml and <0.38 pg/ml, respectively. With these two cut-points, patients in the total cohort were stratified into low-, intermediate-, and high-risk groups for Aβ+. The high-risk group exhibited the steepest annual decline in both MoCA (-2.382) and MMSE (-1.580) scores; while the low-risk group demonstrated relatively stable cognitive performance over time. By applying a k-means clustering analysis, we identified three clusters of participants representing stable, slow, and fast progressors. Fast progressors exhibit the most pronounced cognitive decline and the highest pTau217 levels.

**DISCUSSION:** Plasma pTau217 demonstrates high diagnostic accuracy and effectively predicts cognitive decline even in individuals without known Aβ+ status, potentially reducing reliance on amyloid PET confirmation. Our findings reinforce the clinical utility of plasma pTau217 as a prognostic biomarker for AD progression, offering a practical, non-invasive tool for assessing cognitive decline in real-world Southeast Asian memory clinics.

## 1. BACKGROUND

Blood-based biomarkers have revolutionized Alzheimer’s disease (AD) research, offering a less invasive and cost-effective alternative to traditional diagnostic methods like cerebrospinal fluid (CSF) analysis and PET imaging. These biomarkers enable the detection of key pathological hallmarks of AD, such as amyloid-beta plaques and tau protein tangles, through simple blood tests. Among the most promising biomarkers are phosphorylated tau (pTau) species, including pTau181 and pTau217, which have demonstrated high accuracy in identifying amyloid (Aβ) pathology in AD dementia, MCI and cognitively unimpaired individuals(1–4). Studies have shown that plasma pTau217 has the largest fold-change compared to pTau181 in Aβ-positive vs Aβ-negative individuals(2,5,6), and its levels showed an annual increase only in Aβ-positive individuals, with the greatest change in Aβ-positive tau-positive individuals(2,3). The longitudinal increase in pTau217 levels also associates with cognitive decline and atrophy in Aβ-positive individuals(2). By accurately detecting amyloid pathology, plasma pTau217 has the potential in identifying candidates for amyloid-targeting therapies, and monitoring treatment response during trials.

While plasma pTau217 has garnered significant attention due to its promising results in AD research, its application in real-world settings remains limited, particularly in diverse populations such as those in Asia. Current guidelines for plasma pTau217 is to have 2 cutoffs or a 3-range approach that has demonstrated high negative and positive (>90%) concordance with Aβ status, with approximately 20% of individuals in an intermediate zone that would require confirmatory CSF or PET(7,8). Evaluating diagnostic performance of plasma pTau217 and proposing appropriate cut-off values across diverse populations are essential steps in ensuring its broader clinical applicability and reliability, especially in areas where access to expensive amyloid PET scans and invasive CSF studies are limited. Addressing this gap is crucial to understanding the biomarker’s performance across diverse racial and ethnic groups and improving cost-effectiveness in dementia diagnosis and treatment.

In this study, we evaluated the performance of plasma ptau217 using a commercially available assay with global cognition in a longitudinal Southeast Asian tertiary memory clinic cohort and report reference ranges and cut-offs of plasma pTau217 levels that correspond to a low-, intermediate, or high risk of having a positive amyloid status. Additionally, to investigate patient heterogeneity within diagnostic groups as per a real-world tertiary clinic setting, we applied k-means clustering analysis, an unsupervised machine learning algorithm, to categorize patients into three distinct clusters based on cognitive performance profiles and corresponding plasma pTau217 levels; which provides insights into patterns that may not be immediately evident through traditional analysis.

## 2. METHODS

### 2.1 Study participants

A total of 291 participants [68 healthy controls (HC), 100 mild cognitive impairment (MCI), 86 AD, 37 non-AD dementia] were recruited from specialist memory clinics at the National Neuroscience Institute in Singapore and provided written informed consent through study protocols approved by Singapore Health Services Centralized Institutional Review Board (CIRB). All participants underwent a standardized examination that included a medical evaluation, clinical interview and cognitive screening tests [Mini-Mental State Examination (MMSE) and Montreal Cognitive Assessment (MoCA)] at baseline and annually for 4 years. Participants with MCI and AD patients at a milder dementia stage also undergo more comprehensive evaluation with full neuropsychological assessments (e.g. Colour Trails Test I & II, Repeatable Battery for the Assessment of Neuropsychological Status, and Boston Naming Test) and frailty assessments (e.g. Global Physical Activity Questionnaire, FRAIL scale, Hand grip strength and Clinical Frailty Scale). Diagnosis of different disease groups were made by neurologists specialized in dementia according to international clinical diagnostic criteria(9–14). The non-AD dementia group comprised of frontotemporal dementia, primary progressive aphasia, dementia with Lewy bodies and vascular dementia. Healthy controls were recruited from the community and were free of significant neurological, psychiatric or systemic disease. Apolipoprotein E (*APOE*) genotype was determined in all subjects as described previously(15).

Amyloid status was determined in a subset of patients (n=40) using either Aβ PET imaging with [18F]Flutemetamol or CSF tau/Aβ42 ratio (>0.52)(16). This group consisted of 16 Aβ+ (4 MCI, 12 AD) and 24 Aβ-(14 MCI, 1 HC, 9 frontotemporal lobar degeneration, FTLD) cases. Amyloid PET data were collected in 3D mode with data corrections for 20 minutes following brain CT scans. PET images were reconstructed using iterative methods with CT data used for attenuation correction. A nuclear medicine specialist visually assessed PET images on an Advantage Workstation, with the intensity scale set at 90% for the pons, which served as a reference region. Amyloid deposits in key brain regions, including the frontal, parietal, and temporal lobes, precuneus/posterior cingulate, and striatum, were evaluated. Patients with identified amyloid deposition in these regions were classified as amyloid-positive.

### 2.2 Plasma sampling and analysis

EDTA blood was centrifuged at 1,800 g for 10mins within 1 hour after collection. Plasma was aliquoted and stored at -80 °C until use. Plasma pTau181 was measured using Advantage V2.1 kit) and pTau217 was measured using ALZpath SIMOA p-tau217 Advantage PLUS kits (Quanterix, MA USA) on the HDX Analyzer, according to the manufacturer’s protocol. Inter- and intra-assay coefficients of variation were <15%.

### 2.3 Statistical analysis

Descriptive statistics were used to summarize baseline demographic, clinical, and biomarker characteristics. Continuous variables were assessed for normality and described using mean ± standard deviation or median with interquartile range, as appropriate. Categorical variables were summarized using frequencies and percentages. Comparisons of continuous variables across diagnostic and amyloid groups, as well as across cognitive trajectory clusters, were conducted using the Kruskal–Wallis test. Chi-square or Fisher’s exact tests, where appropriate, were used to compare categorical variables among the above-mentioned groups. Plasma pTau217 and pTau181 were natural log-transformed to reduce skewness prior to subsequent analyses.

Discriminative performance of plasma pTau217 and pTau181 for AD diagnosis and amyloid positivity was evaluated using receiver operating characteristic (ROC) curve analysis, and DeLong’s test was used to compare AUCs. Fold-differences in mean biomarker levels between groups were also calculated.

Cut-points for pTau217 were determined based on thresholds corresponding to 95% sensitivity and 95% specificity from the ROC curve for amyloid positivity. The 95% sensitivity threshold was used to define the lower bound, and the 95% specificity threshold defined the upper bound. Participants were then stratified into three risk categories: low (pTau217 < sensitivity threshold), intermediate (between the sensitivity and specificity thresholds), and high (pTau217 > specificity threshold) for subsequent analyses.

To assess the relationship between pTau217 levels and longitudinal cognitive decline, we fitted linear mixed-effects models (LMMs) with MoCA and MMSE as dependent variables. Fixed effects included time (years), pTau217 risk group (low, intermediate, high), and the interaction term (time × pTau217 group), adjusting for age, gender, education, and diagnosis. Model diagnostics were checked to assess underlying assumptions, and estimated slopes were used to quantify group-specific rates of cognitive change.

To explore data-driven subtypes, k-means clustering was performed using demographic variables (age, gender, education, *APOE4*), baseline cognitive scores (MMSE, MoCA), pTau217 levels, and a cognitive progression score. Progression was defined as the most negative annualized change in MoCA over four years. Participants with at least two non-missing MoCA observations were included. Missing data in education and *APOE4* were imputed using the MissForest method. Cognitive trajectories across the clusters were further analyzed using an additional LMM with fixed effects for time, cluster, and their interaction.

All statistical analyses were performed using R software (R Core Team (2024); R: A Language and Environment for Statistical Computing. R Foundation for Statistical Computing, Vienna, Austria. https://www.R-project.org) with a two-sided p-value < 0.05 considered statistically significant.

## 3. RESULTS

### 3.1 Baseline characteristics

Demographic information and clinical characteristics of the total participants and subset of participants with amyloid status are presented in Table 1. A total of 291 participants [mean (SD) age= 63.5 (8.9) years; 50% males; 91% Han Chinese ethnicity] were included in the study. Overall, ptau217 correlated with age (rs= 0.126, p=0.032), plasma pTau181 (rs= 0.740, p<0.001), MMSE (rs= -0.543, p<0.001) and MoCA (rs= -0.551, p<0.001) at baseline, but not with gender (p= 0.275).

**Table 1.** Demographic and clinical characteristics of all subjects at baseline.

| Characteristic<sup>a</sup> | HC | MCI | AD | Non-AD | p value<sup>b</sup> | Aβ positive<sup>c</sup> | Aβ negative | p value<sup>d</sup> |
| --- | --- | --- | --- | --- | --- | --- | --- | --- |
| Subjects, n | 68 | 100 | 86 | 37 |  | 16 | 24 |  |
| Age, years | 62.1 ± 9.6 | 64.3 ± 8.7 | 64.4 ± 8.7 | 61.9 ± 8.2 | 0.311 | 60.8 ± 6.8 | 61.4 ± 6.3 | 0.759 |
| Male sex, n (%) | 40 (59%) | 51 (51%) | 40 (47%) | 14 (40%) | 0.201 | 7 (44%) | 15 (62%) | 0.243 |
| Race (C:M:I:O) | 65:0:2:1 | 90:2:4:4 | 79:4:1:2 | 32:1:3:1 | 0.476 | 14:0:0:2 | 19:1:3:1 | 0.294 |
| Education, years | 15.9 ± 3.5 | 12.8 ± 4.0 | 10.3 ± 4.3 | 9.3 ± 5.1 | <0.001 | 12.5 ± 4.2 | 11.8 ± 4.8 | 0.653 |
| APOE4 carriers | 13 (21%) | 21 (24%) | 28 (39%) | 4 (13%) | 0.018 | 4 (27%) | 11 (73%) | 0.900 |
| MMSE | 29.0 ± 1.4 | 27.7 ± 2.4 | 18.9 ± 6.4 | 21.4 ± 7.4 | <0.001 | 22.0 ± 5.9 | 26.7 ± 4.2 | 0.007 |
| MoCA | 27.4 ± 1.8 | 26.0 ± 3.0 | 14.6 ± 7.1 | 18.7 ± 7.8 | <0.001 | 18.1 ± 7.5 | 25.1 ± 5.9 | 0.003 |
| Plasma pTau217 (pg/mL) | 0.295 ± 0.107 | 0.500 ± 0.522 | 1.433 ± 0.846 | 0.273 ± 0.143 | <0.001 | 1.338 ± 0.649 | 0.285 ± 0.181 | <0.001 |
| Plasma pTau181 (pg/mL) | 18.984 ± 5.111 | 22.419 ± 8.916 | 44.976 ± 19.037 | 23.332 ± 16.125 | <0.001 | 43.351 ± 17.016 | 19.073 ± 8.002 | <0.001 |

Abbreviations: HC, Healthy Control; MCI, Mild Cognitive Impairment; AD, Alzheimer's disease; Race: C=Chinese, M=Malay, I=Indian, , O=Others; MMSE, Mini Mental State Examination; MoCA, Montreal Cognitive Assessment.
<sup>a</sup> Continuous variables reported as mean ± standard deviation; Categorical variables reported as frequency (%).
<sup>b</sup> p value across clinical disease groups.
<sup>c</sup> Out of the 40 patients with amyloid status, 16 Aβ + (4 MCI, 12 AD) and 24 Aβ - (14 MCI, 1 HC, 9 are non-AD dementia).
<sup>d</sup> p value between amyloid-positive and negative groups.

### 3.2 Plasma pTau181 and pTau217 by clinical diagnosis in total cohort

Plasma ptau217 was significantly higher in AD compared to MCI, non-AD dementia and HC (all p<0.001; Figure 1), controlling for age and gender. Plasma ptau217 was significantly higher in MCI vs non-AD dementia (p=0.028) and HC (p=0.030), controlling for age and gender. pTau181 was significantly higher in AD vs MCI, non-AD dementia and HC (all p<0.001), but not significant between MCI vs non-AD dementia (p=1.0) and HC (p=0.350), controlling for age and gender. When comparing performance in differentiating AD vs other diagnostic groups, plasma ptau217 outperformed ptau181 [AUC (95% CI)= 0.942 (0.910, 0.975) vs 0.909 (0.858, 0.961)]. Fold differences between AD and HC means were 4.86 and 2.36 for ptau217 and pTau181, respectively.

**Figure 1.**
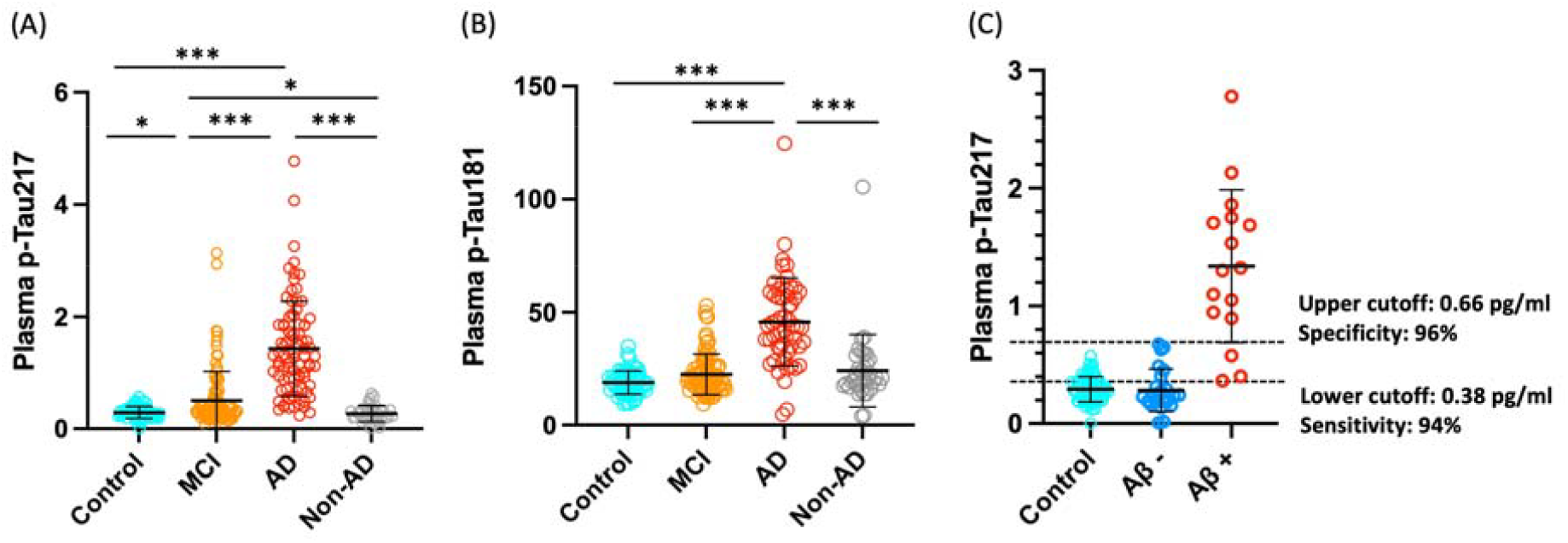
Plasma biomarkers by clinical diagnosis and amyloid status. A) Plasma pTau217 levels across clinical groups; B) Plasma pTau181 across clinical groups; C) Plasma pTau217 levels across healthy controls, amyloid positive and negative groups with two cut-points for pTau217 determined based on thresholds corresponding to 95% sensitivity and 95% specificity for amyloid positivity. *p<0.05, **p<0.01, p values are adjusted for age, gender and multiple comparisons via Bonferroni. MCI, Mild Cognitive Impairment; AD, Alzheimer’s disease.

### 3.3 Plasma pTau181 and ptau217 in Aβ−and Aβ+ participants

Among patients with brain amyloid status (n=40; 16 Aβ+, 24 Aβ-), pTau217 identified Aβ+ with AUC= 0.961 (95% CI= 0.909-1.000, sensitivity= 0.81, specificity=1.00) vs pTau181 with AUC= 0.917 (95% CI= 0.825-1.000, sensitivity= 0.94, specificity=0.83) (P _difference_ = 0.230). Fold differences between Aβ+ and Aβ+-means were 4.69 and 2.27 for ptau217 and pTau181, respectively. The addition of age, gender and *APOE4* status to pTau217 increased the AUC= 0.982 (95% CI= 0.948-1.000) and for pTau181 (AUC= 0.935, 95% CI= 0.863-1.000) in differentiating Aβ positivity (P_difference_ = 0.164). When comparing each biomarker model with and without the addition of age, gender, and *APOE4*, they did not significantly improve the AUC for pTau217 (P_difference_ = 0.376) or for pTau181 (p = 0.621). These findings suggest that while both biomarkers show high discriminative ability for Aβ positivity, the differences between them, and the incremental gain from adding demographic and genetic covariates, are not statistically significant.

### 3.4 Definition of pTau217 cut-point to detect cerebral amyloidosis

As pTau217 has superior accuracy to ptau181 in identifying Aβ+, we derived pTau217 cut-offs for further analysis. Using a two cut-point approach, 95% sensitivity and specificity for Aβ+ corresponded to pTau217 >0.66 pg/ml and <0.38 pg/ml, respectively. With these two cut-points for pTau217, patients in the total cohort were stratified into 155 (53.3%) low-, 48 (16.5%) intermediate-, and 88 (30.2%) high-risk groups for Aβ+.

### 3.4 Association between baseline pTau217 and cognitive performance over 4 years

With these two cut-points for pTau217, patients in the total cohort were stratified into 155 low-, 48 intermediate-, and 88 high-risk groups for Aβ+. The results from the linear mixed-effects model indicate significant differences in the progression of cognitive decline, as measured by MMSE and MoCA, across different pTau217 groups (Table 2, Figure 2) controlling for age, gender and education. Based on the estimated slopes for each group, the high-risk group (>0.66 pg/ml) exhibited the steepest decline in both MMSE (-1.580) and MoCA (-2.382), suggesting a more rapid cognitive deterioration. In contrast, the low-risk group (<0.38 pg/ml) demonstrated relatively stable cognitive performance over time, with a slope estimate close to zero for both MMSE (0.063) and MoCA (0.285). The Intermediate group showed a moderate decline, falling between the low- and intermediate-groups. MMSE and MoCA were chosen for analysis as they are the most commonly used tools for global cognition in a real-world memory clinic, making these results more clinically applicable.

**Table 2.**
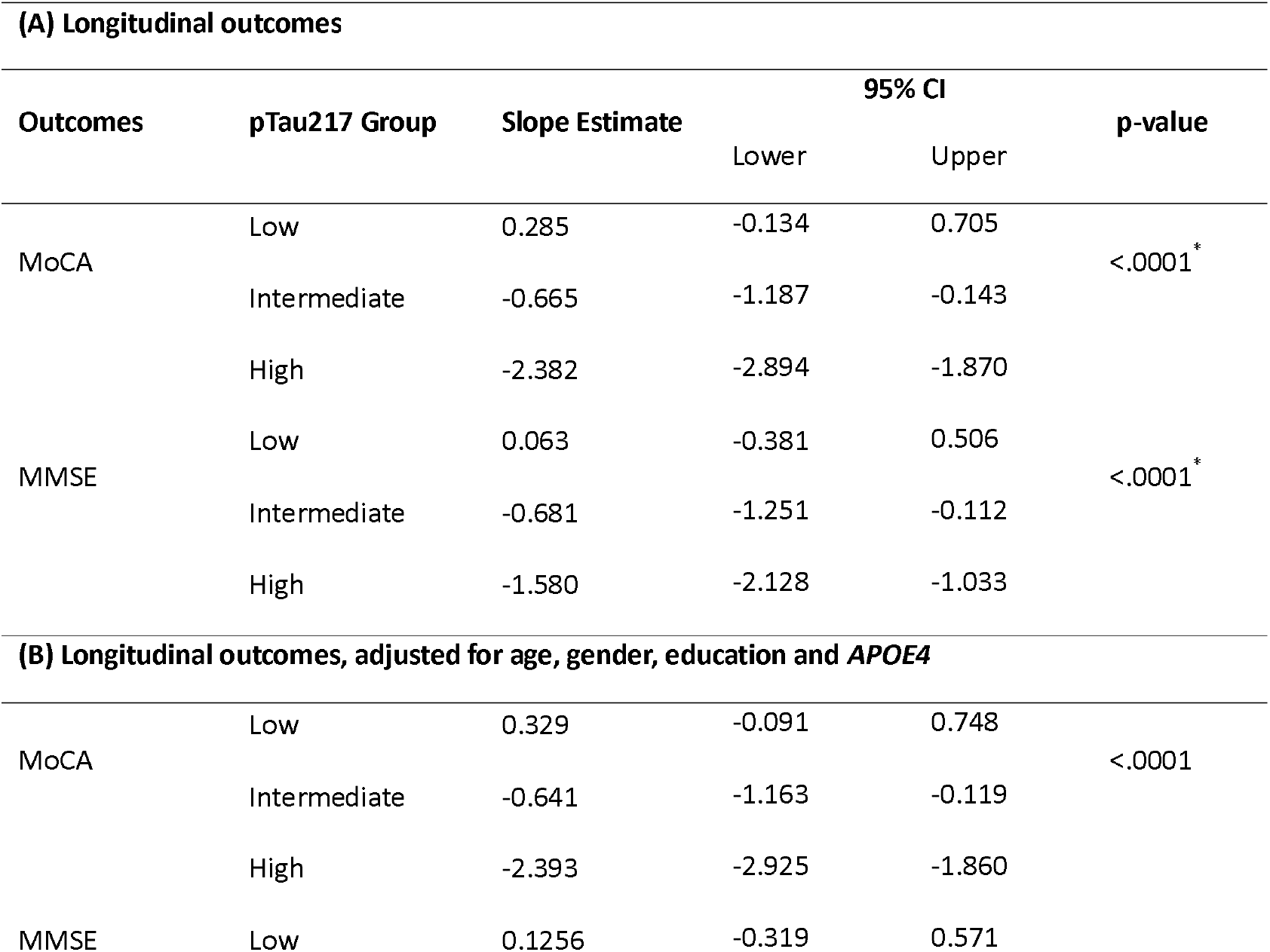
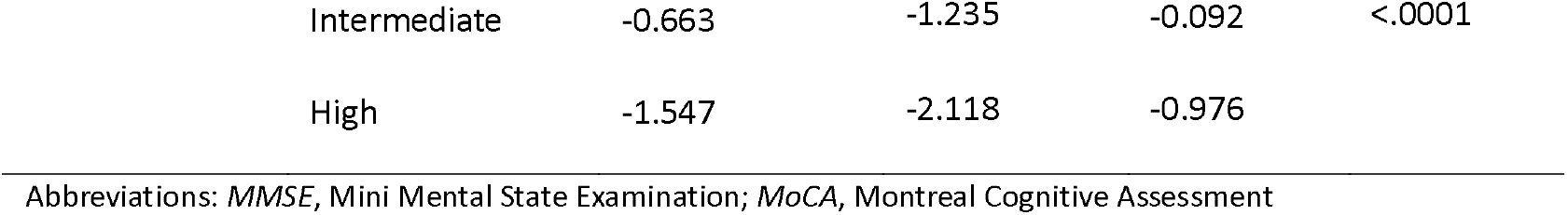
Comparison of MMSE and MoCA progression rates among three pTau217 groups, adjusted for age, gender, and education.

**Figure 2.**
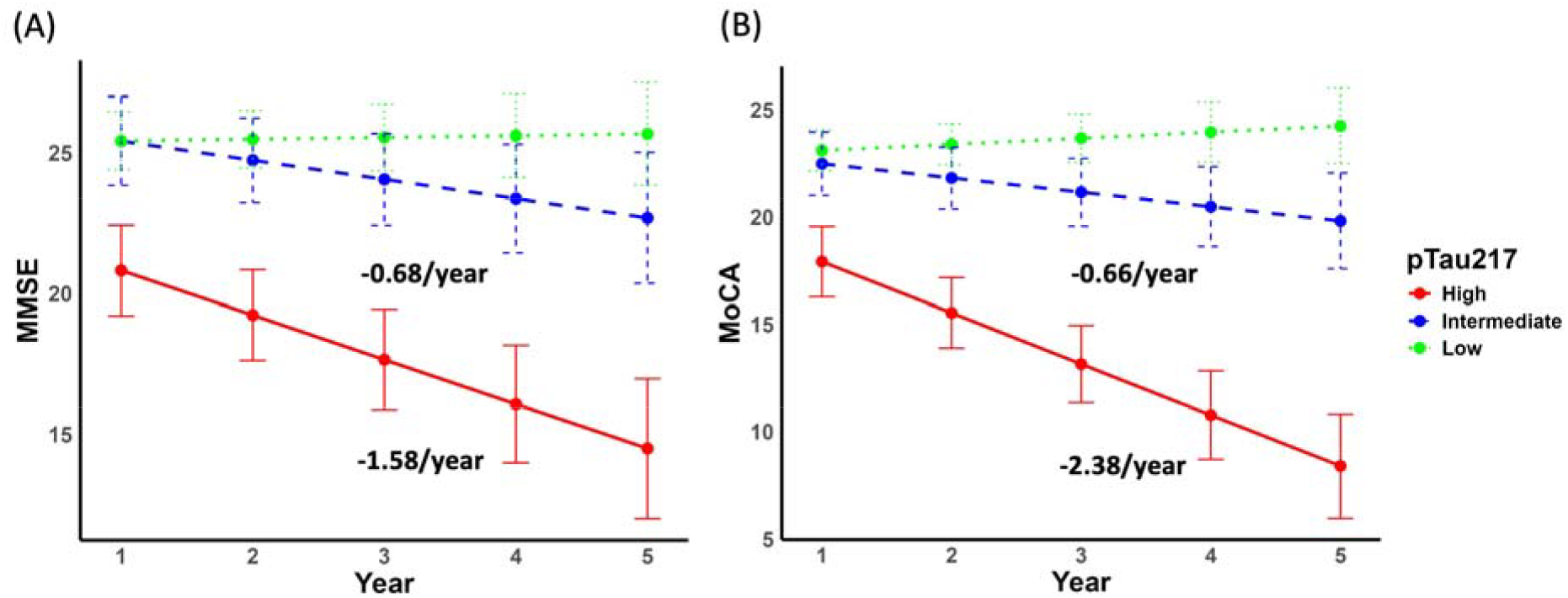
Associations between baseline plasma ptau217 risk groups with longitudinal cognitive performance over 4 years. Longitudinal trajectories of (A) MMSE and (B) MoCA scores over 4 years for the three pTau217 groups (low, Intermediate, and high). The high-risk group (>0.66 pg/ml) exhibited the steepest decline in both MMSE and MoCA, while the low-risk group (<0.38 pg/ml) demonstrated relatively stable cognitive performance over 4 years. The error bars represent 95% confidence intervals.

### 3.5 Cluster analysis and association with pTau217 levels

To elucidate patient heterogeneity across diagnostic groups as frequently encountered in a real-world memory clinic, we performed k-means clustering to classify participants with longitudinal data into three distinct subtypes based on baseline demographic characteristics (age, years of education), cognitive performance (MMSE and MoCA scores), plasma pTau217 levels, and cognitive progression over a four-year period. The resulting clusters were labeled as stable (Cluster 1), slow progressors (Cluster 2), and fast progressors (Cluster 3), reflecting their cognitive decline trajectories and providing insights into the alignment of plasma pTau217 with disease progression and clinical outcomes.

Cluster characteristics and cognitive progression scores are summarized in Table 3. Cluster 3 (fast progressors) exhibited the most rapid cognitive decline, with a median progression score of -3.0 on the MoCA, indicating significantly greater deterioration compared to Cluster 2 (slow progressors, -1.33) and Cluster 1 (stable, 0). This cluster was further distinguished by lower baseline cognitive performance (median MMSE = 17, MoCA = 10), younger age (median = 59 years), fewer years of education (median = 10.0 years), and elevated plasma pTau217 levels (median = 1.69 pg/ml). In contrast, Cluster 1 (stable) showed minimal cognitive decline, while Cluster 2 (slow progressors) displayed moderate progression. Data from the clustering analysis performed serves to reinforce our earlier results.

**Table 3.**
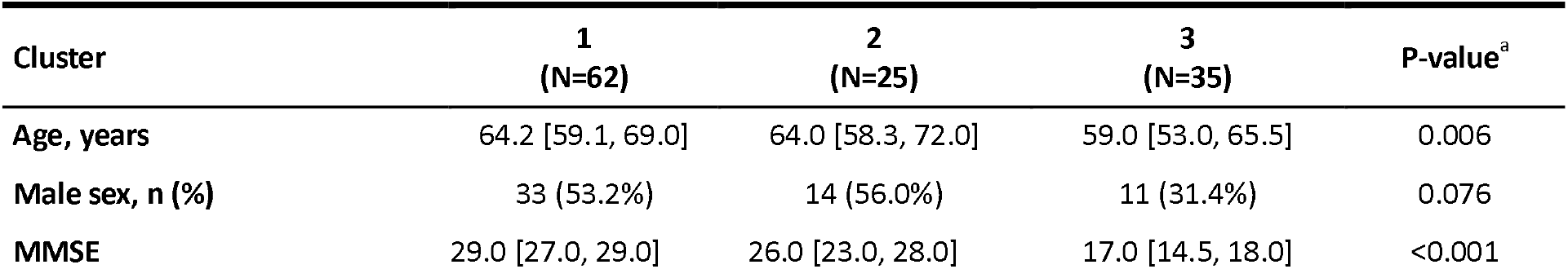
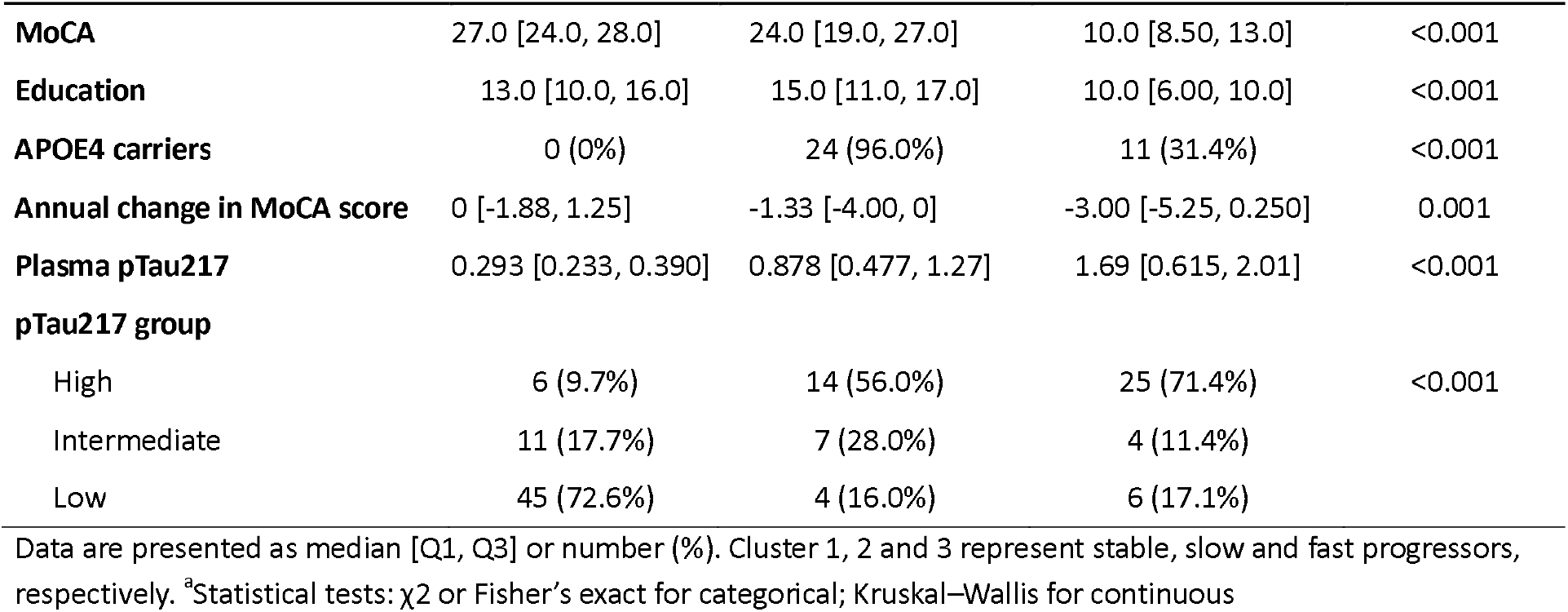
Baseline characteristics and progression across three cognitive trajectory groups identified by cluster analysis.

## 4. DISCUSSION

In this study, we evaluated the performance of plasma pTau217 using a commercially available assay (ALZpath SIMOA) as a predictor of amyloid pathology and cognitive decline in an Asian tertiary memory clinic. We established cutpoints with robust discriminative performance of pTau217 for positive vs negative Aβ status, superior to pTau181 (consistent in other large cohorts(3,17)). We presented two cutpoints (pTau217 >0.66 pg/ml and <0.38 pg/ml) with 95% sensitivity and 95% specificity for Aβ+. Using these cut points, 16.8% of participants fell into the intermediate category, aligning with the recommendations set forth by the Global CEO Initiative on Alzheimer’s Disease to minimize the number of individuals that must undergo additional testing(7). Moreover, individuals in the high-risk group (>0.66 pg/ml) exhibited a greater rate of cognitive decline (annual decline rate of -1.580 for MMSE and -2.382 for MoCA). Importantly, we conducted k-means cluster analysis to classify participants into three distinct subtypes (slow, stable and fast progressors) based on baseline characteristics and cognitive progression over a four-year period. We showed that fast progressors had the highest pTau217 levels, youngest age and worst cognition at baseline. Results from the clustering analysis reinforce our findings, providing valuable insights into how plasma pTau217 aligns with disease trajectories and clinical outcomes across different patient groups, even in MCI and AD patients without amyloid PET confirmation.

First, we observed higher ptau217 in AD compared to MCI, non-AD dementia and HC, and MCI vs non-AD dementia and HC. Importantly, we demonstrate that the fold change for pTau217 between clinically diagnosed AD (without evaluation of amyloid status) and HC in the total cohort was larger than that for pTau181, similar to the higher fold change for pTau217 seen in the subset of patients with amyloid status (4.86 vs 4.69 for Aβ+, respectively), as well as those reported in other cohorts (fold changes range 1.5-4.42 pg/ml(6,18–20)). Our findings also demonstrated the robust discriminative performance of plasma pTau217 for positive vs negative Aβ status with an AUC of 0.957, comparable to other studies(21–24). Our findings of higher fold-change and accuracy for ptau217 vs ptau181 is in line with prior studies where plasma pTau217 has been shown to differentiate pathologically diagnosed AD from non-AD with significantly higher accuracy than pTau181(17,25). Only plasma pTau217 was able to differentiate between all levels of AD neuropathological change (none, low, intermediate, and high), while pTau181 only differentiated between intermediate and high AD neuropathology(25). Plasma pTau217 outperformed plasma pTau181 when discriminating clinically diagnosed and pathology-confirmed AD from frontotemporal lobar degeneration(5), while another recent study showed the potential of plasma ptau217 in detecting AD copathology in non-AD neurodegenerative syndromes(26).

From a subset of patients with amyloid status, we derived two cutpoints (pTau217 >0.66 pg/ml and <0.38 pg/ml) with 95% sensitivity and specificity for Aβ+. These cutpoints are similar to the ones reported by Ashton et al(3) (>0.63pg/ml and <0.40 pg/ml) and Mammel et al(22) (>0.63pg/ml and <0.34 pg/ml), both using the ALZpath pTau217 assay. Despite differences in ethnicity, the mean age of participants in their study and ours are similar (mean age of 64 years), which is younger than other cohorts reporting ALZpath cutoffs(6,20). The lower cutoffs (<0.23-0.386pg/ml) reported in other studies involving older cohorts (mean age in the 70s) using ALZpath are similar to our study, however they reported significantly different upper cutoffs (Figdore et al(20). at 0.811 pg/ml; Lehmann et al(6) at 0.80 pg/ml). The reason for this discrepancy remains unclear, but maybe due to demographic difference in study populations or differences in the methodology for determining amyloid status. These findings underscore the importance of additional studies such as ours to reevaluate and refine cut points for the pTau217 assay in other ethnic groups. Such efforts are crucial to ensure reliability and consistency before widespread clinical implementation, particularly across diverse populations. With these two cut-points, patients in the total cohort were stratified into low-, intermediate-, and high-risk groups for Aβ+. The high-risk group (>0.66 pg/ml) exhibited the steepest annual decline in both MMSE (-1.580) and MoCA (-2.382), suggesting a more rapid cognitive deterioration. In contrast, the low-risk group (<0.38 pg/ml) demonstrated relatively stable cognitive performance over time, with a slope estimate close to zero for both MMSE and MoCA. The Intermediate group showed a moderate decline, falling between the low- and intermediate-groups.

K-means clustering identifies subgroups within a population by grouping individuals with similar characteristics, enabling a data-driven approach to explore relationships between biomarkers, cognitive decline, and disease progression without requiring predefined clinical thresholds. We performed additional clustering analysis in our cohort and were able to distinctly categorize participants into stable, slow, and fast progressors over a 4-year period, where fast progressors exhibited the highest median pTau217 levels (1.69 pg/ml), followed by slow (0.878 pg/ml) and stable (0.293 pg/ml) groups. The clustering results provided further validation of the rates of cognitive decline within each subgroup through rigorous statistical testing and reinforced our earlier observations, important when evaluating plasma pTau217 with clinical outcomes within heterogenous diagnostic groups reflective of a real-world memory clinic.

Although plasma pTau217 has shown exceptional accuracy in research cohorts, the majority of studies to date have been conducted in Western populations, leaving a significant gap in understanding its performance across diverse ethnic groups. To bridge this gap, we evaluated the performance of plasma pTau217 in Singapore to assess its applicability in this underrepresented population. A study from Thailand applied a Bayesian approach with prior probabilities to evaluate plasma pTau217 in memory clinic patients and demonstrated high diagnostic accuracy(27). Another study from Taiwan reported high diagnostic accuracy of pTau217 in memory clinic patients from Taiwan and Korea(23) and classified participants into low-, intermediate-, and high-risk groups based on model-derived probabilities. Both studies reinforce the diagnostic potential of plasma pTau217 in real-world settings, but our study provides important additional insights into disease progression (including magnitude of cognitive decline annually), prognostic utility, and practical cutpoints for clinical implementation. Reporting pTau217 studies in real-world settings is particularly important because such environments reflect the complexities and variability seen in routine clinical practice.

The main limitation is the inclusion of a smaller sample size of cases with known amyloid status confirmed via amyloid PET or CSF testing. However, the reasonably large MCI and AD group recruited from a single specialized center, reflective of the diagnostic make-up of real-world memory clinics, underscores the validity and high diagnostic accuracy of plasma pTau217 even in patients without known amyloid status but diagnosed clinically to have MCI or AD, potentially sparing patients from invasive CSF testing or costly amyloid PET imaging. Importantly, we were also able to derive a 2-point cut-off from our cohort that was very similar to internationally published cut-offs(3,22). Another strength of this study includes our longitudinal clinical follow-up over a period of 4 years. There are limited studies on ptau217 in Asian memory clinics(23,27), most of which are cross-sectional in nature. Apart from reporting a cutpoint of pTau217 >0.66 pg/ml for amyloid positivity with a 95% sensitivity and specificity, we also reported elevated level (1.69 pg/ml) in fast progressors, as identified through cluster analysis. Patients within this group not only exhibit the highest plasma pTau217 levels but also experience the steepest trajectory of disease progression. This finding suggests that higher pTau217 levels may indicate more aggressive neurodegeneration, emphasizing its utility in stratifying patients for closer monitoring and targeted interventions.

In conclusion, our study adds to the growing body of evidence supporting the utility of plasma pTau217 as a reliable diagnostic and prognostic biomarker for AD, especially in Southeast Asia. The robust predictive performance of plasma pTau217 cutpoints underscores its potential in guiding clinical decisions, reducing reliance on invasive and expensive confirmatory tests, and prioritizing early intervention strategies. Future research on plasma pTau217 should focus on validating the derived cut-off values across different age groups and ethnicities to ensure consistency and accuracy and expanding studies to more real-world settings, such as specialized memory clinics, will provide valuable insights into its performance in varied clinical contexts. These findings are instrumental in ensuring plasma pTau217 becomes a reliable, scalable biomarker for early detection, monitoring, and tailored care within the Alzheimer’s disease continuum.

## Data Availability

Anonymised data that support the findings of this study are available from the corresponding author upon reasonable request.

## Ethics approval and consent to participate

All participants provided written informed consent through study protocols approved by Singapore Health Services Centralized Institutional Review Board (CIRB).

## Authors’ contributions

ASLN and YJT designed the study. YJT, NIT, LSSL, SHLL,LCST, HJC, KPN, SH, SKST, and ASLN acquired the data relevant for the study. YJT, RM, and SES analysed, interpreted the data, and drafted the manuscript. All authors contributed to the writing and revisions of the paper and approved the final version.

## Acknowledgements

The authors sincerely thank all participants and research staff, especially Karie Chua and Valerie Teh from the Department of Research, National Neuroscience Institute, Singapore, for their invaluable contribution to this study. We also extend our appreciation to Regina Chen and Yi Hao Nah from the Department of Research, National Neuroscience Institute, Singapore, for their efforts in recruiting healthy controls utilized in our research. We thank Dr Xie Wanying from the Department of Nuclear Medicine, Singapore General Hospital, Singapore, for her invaluable contribution in the field of amyloid PET imaging.

